# Residual Symptoms and Quality of Life in Female Patients with Hypothyroidism and Cardiovascular Disease

**DOI:** 10.64898/2026.10.30.26366962

**Authors:** Corinthia S. Gonzales, Emma Theisen, Alexandra Sarau, Joohye Kim, Daniel C. Landek, Antonio C. Bianco, Annabelle Santos Volgman

**Author notes:** **Corresponding Author:** Annabelle Santos Volgman, MD, FACC, FAHA, McMullan-Eybel Endowed Chair for Excellence in Clinical Cardiology Professor of Medicine, Rush College of Medicine, RUSH University Medical Center, 1725 W Harrison St, Suite 1159, Chicago, IL 60612.

## Abstract

**Background:** Current clinical management of patients with hypothyroidism emphasizes thyroid hormone replacement at doses that normalize serum thyroid-stimulating hormone (TSH) levels. However, biochemical correction does not always translate into symptom resolution or restored health-related quality of life (QoL). We evaluated whether higher TSH levels within the reference range would be associated with worse Thyroid-Related Patient-Reported Outcome (ThyPRO)-39 scores, reflecting greater symptom burden.

**Objective:** The objective of this study was to assess residual symptoms and health-related quality of life in women with normal TSH levels in patients with cardiovascular disease being treated for hypothyroidism.

**Materials and methods:** We prescreened 96 participants who completed study assessments. Following the TSH obtained at the study visit, 14 participants were excluded due to TSH values falling outside the predefined euthyroid range (0.35-4.94 μIU/mL), resulting in a final cohort of 82 women. We also investigated whether comorbidities contributed to symptom burden.

**Results:** Although mean composite scores were numerically higher in the upper TSH quartiles, neither multivariable regression nor ordered trend testing demonstrated a significant association between increasing TSH and overall symptom burden (Jonckheere-Terpstra p = .333). Notably, major depressive disorder (MDD) (β=16.93, 95% CI 8.22–25.65; p<.001) and hypertension (HTN) (β=12.45, 95% CI 2.24–22.66; p=.018) were independently associated with greater Thy-PRO-39 scores.

**Conclusions:** MDD and HTN were independently associated with higher Thy-PRO-39 scores, suggesting that comorbidities may be more strongly related to persistent symptoms than modest variation in TSH within the euthyroid range. Our results suggest that in patients with hypothyroidism and cardiovascular disease, comorbidities may be particularly important when interpreting residual symptoms in biochemically euthyroid patients.

## Introduction

Hypothyroidism is a common multisystem disorder characterized by a deficiency of thyroid hormones, with wide-ranging effects mainly on metabolic, neurologic, and reproductive systems.^1^ Thyroid hormone also plays an important cardiovascular role by affecting cardiomyocytes, vascular smooth muscle, and endothelial cells; hypothyroidism is associated with alterations in lipid metabolism, blood pressure regulation, and cardiac contractility.^2,3^

Current clinical management of patients with hypothyroidism emphasizes replacement of thyroid hormone at doses that normalize serum thyroid-stimulating hormone (TSH) levels, which also reverses many of these abnormalities.^4^ However, biochemical correction does not uniformly translate into symptom resolution or restoration of health-related quality of life (QoL). Indeed, despite achievement of biochemical euthyroidism with levothyroxine (LT4) therapy, a substantial proportion of patients with hypothyroidism report persistent symptoms, including fatigue, cognitive impairment, mood disturbances, and reduced overall well-being.^5^ Patient dissatisfaction with LT4 therapy remains common, with individuals expressing ongoing symptom burden and a desire for improved treatment strategies.^6^ A meta-analysis of randomized controlled trials has further demonstrated that LT4 supplementation does not consistently result in significant improvements in QoL measures, even when serum TSH levels are normalized.^7^ Proposed explanations for persistent symptoms include incomplete normalization of thyroid hormone signaling at the tissue level, autoimmune-related mechanisms, and the contribution of coexisting systemic or psychosocial comorbidities.^8^ This scenario highlights a disconnect between biochemical targets and patient-reported outcomes in the management of hypothyroidism.

Given this gap, patient-reported outcome measures have become increasingly important tools for assessing the impact of thyroid disease and its treatment. The Thyroid-Related Patient-Reported Outcome (ThyPRO) questionnaire was developed as a disease-specific instrument to assess QoL in patients with thyroid disorders and has demonstrated strong validity, reliability, and responsiveness to treatment effects.^9^ A shortened 39-item version (ThyPRO-39) (Supplemental Figure) was subsequently validated and shown to perform comparably to the original instrument while reducing respondent burden.^10^ The ThyPRO has also demonstrated cross-cultural and cross-linguistic validity, supporting its use across diverse patient populations.^11^ Studies using the ThyPRO have continued to yield mixed findings regarding the relationship between LT4 treatment, biochemical improvement, and QoL. Winther et al. reported significant improvement across multiple ThyPRO domains during the first six months of LT4 treatment in autoimmune hypothyroidism. In contrast, Stott et al. found that lowering TSH with LT4 in older adults with subclinical hypothyroidism did not significantly improve QoL.^12,13^

It is well known that women are disproportionately affected by hypothyroidism and are more likely to report persistent symptoms while appropriately treated, particularly symptoms that overlap with cardiovascular complaints such as fatigue, exercise intolerance, and palpitations.^6,14^ These nonspecific symptoms frequently prompt evaluation in specialty settings, where distinguishing thyroid-related symptom burden from other contributors remains clinically challenging. A study of 2203 adult patients found that suboptimal levothyroxine treatment was associated with increases in mean BP and BP variability, which may be important in adequate treatment of hypertension.^14^

Understanding the relationship between biochemical thyroid control and patient-reported QoL in this population is therefore of particular relevance to real-world clinical care. Therefore, the objective of this study was to assess residual symptoms and health-related quality of life in women with hypothyroidism being treated in an outpatient cardiology office in a major medical center. We evaluated whether higher TSH levels within the reference range would be associated with worse ThyPRO-39 scores, reflecting greater symptom burden. We also investigated whether comorbidities and hypertension contributed to symptom burden.

## Materials and methods

This prospective observational cohort study was conducted at the Rush Heart Center for Women. This study was approved on May 24, 2023, by the Rush University Medical Center Institutional Review Board (IRB Protocol #23010401-Oncore01). Eligible participants were women aged 18 years or older with a prior diagnosis of overt hypothyroidism, receiving stable levothyroxine (LT4) therapy at ≥ 75 μg/day for at least six months, and with documented TSH levels within the institutional reference range (0.35–4.94 μIU/mL) in the preceding 12 months. We excluded patients with TSH, free T4, or T3 values outside the normal range; a history of thyroidectomy; malignant thyroid disease; or an LT4 dose < 75 μg/day. Patients presenting to the Rush Heart Center for Women meeting the inclusion criteria were invited to join the study. Enrollment ended in September 2025.

At study enrollment, participants completed the ThyPRO-39 questionnaire. We also obtained repeat TSH levels at this time. Participants whose contemporaneous TSH values fell outside the institutional reference range were excluded from the final analytic cohort. TSH values were categorized into predefined quartiles: Q1 (0.35–1.00 μIU/mL), Q2 (1.01–2.00 μIU/mL), Q3 (2.01–3.00 μIU/mL), and Q4 (3.01–4.94 μIU/mL). We scored ThyPRO-39 responses as previously reported, with transformed domain scores ranging from 0 to 100 and higher scores indicating greater symptom burden or poorer thyroid-related quality of life. Scores were calculated for the 12 ThyPRO-39 symptom and functional domains. We calculated a composite symptom burden score from the seven well-being and function scales and the single overall quality-of-life item, following published scoring procedures.

Associations between TSH quartiles and ThyPRO composite symptom burden were evaluated using multivariable linear regression. The model was adjusted for clinically relevant covariates including age, body mass index (BMI), vitamin B and D deficiency, major depressive disorder (MDD), hypertension (HTN), and Charlson Comorbidity Index (CCI). Given the hypothesis that symptom burden increases with higher-normal TSH levels, we also evaluated ordered trends across TSH quartiles using the Jonckheere-Terpstra nonparametric trend test.

Exploratory analyses examined associations between continuous TSH concentration and individual ThyPRO-39 domain scores using separate multivariable linear regression models adjusted for the same covariates. False discovery rate correction was performed using the Benjamini-Hochberg method. Associations between TSH and prevalent hypertension were evaluated using multivariable logistic regression, while associations with systolic and diastolic blood pressure were evaluated using separate multivariable linear regression models; these were corrected for age, BMI, and menopausal status. All statistical analyses were performed using R version 4.3.1. Statistical significance was defined as p < 0.05.

## Results

A total of 96 participants were prescreened and completed study assessments. Following the TSH obtained at the study visit, 14 participants were excluded due to TSH values falling outside the predefined euthyroid range (0.35-4.94 μIU/mL), resulting in a final cohort of 82 women. Table 1 presents baseline demographic and clinical characteristics of the study cohort, stratified by TSH quartile. Participants were categorized according to TSH quartile: Q1 (0.35–1.00 μIU/mL, n = 25), Q2 (>1.00–2.00 μIU/mL, n = 31), Q3 (>2.00–3.00 μIU/mL, n = 21), and Q4 (>3.00–4.94 μIU/mL, n = 5). Mean ThyPRO-39 composite scores were 22.9 in Q1, 27.8 in Q2, 27.3 in Q3, and 29.1 in Q4 (Figure 1).

**Figure 1.**
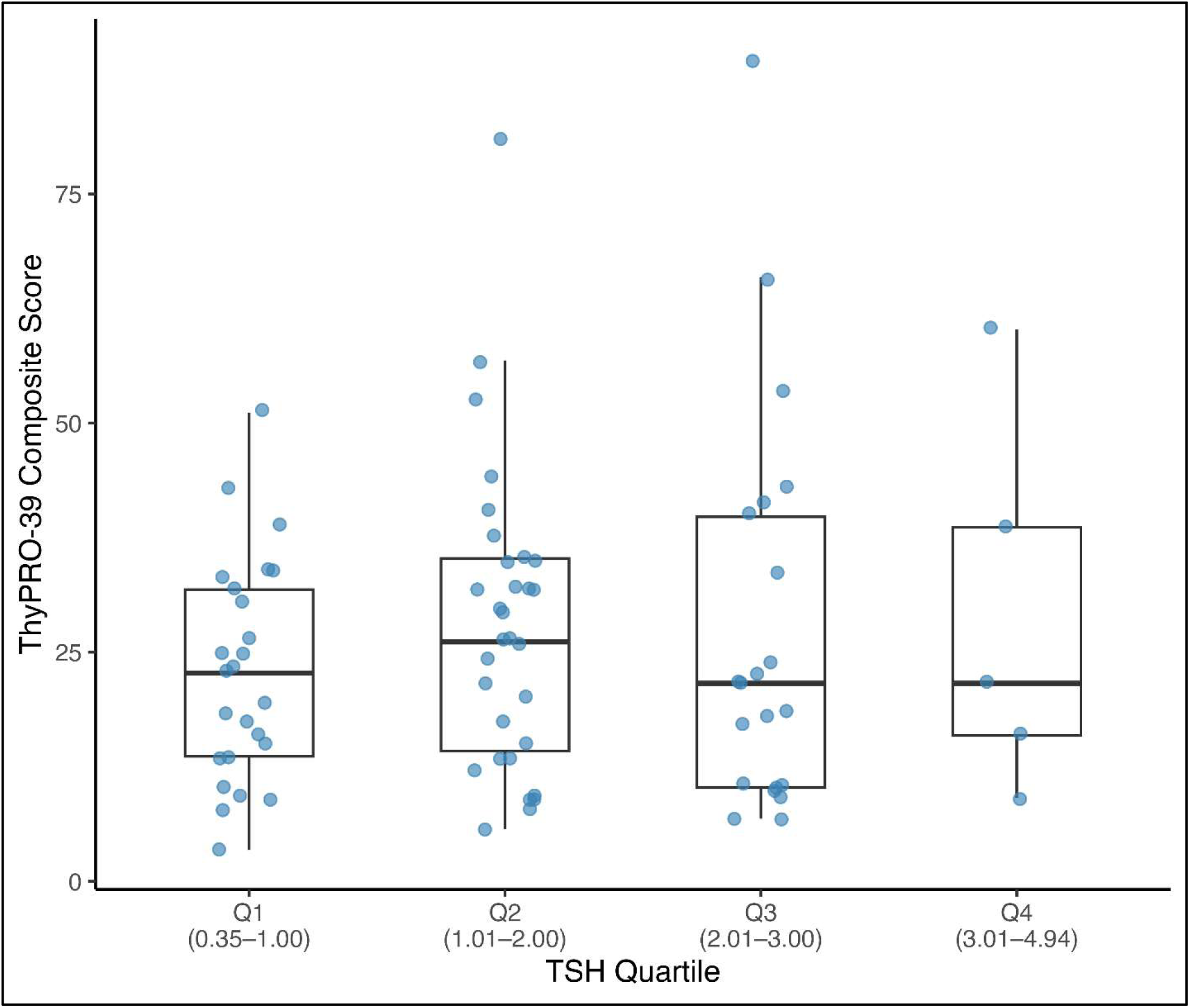
ThyPRO-39 composite score by TSH quartiles.

**Table 1.** Baseline demographic and clinical characteristics of the study cohort, stratified by TSH quartile. Abbreviation – TSH-serum thyroid-stimulating hormone.

| **Characteristic** | **Overall**<br>N = 82<sup>1</sup> | **Q1**<br>N = 25<sup>1</sup> | **Q2**<br>N = 31<sup>1</sup> | **Q3**<br>N = 21<sup>1</sup> | **Q4**<br>N = 5<sup>1</sup> | **p-value**<sup>2</sup> |
|---|---|---|---|---|---|---|
| **Age, years** | 71 ± 12 | 70 ± 13 | 68 ± 11 | 75 ± 10 | 78 ± 9 | 0.11 |
| **BMI, kg/m²** | 29 ± 7 | 29 ± 6 | 30 ± 7 | 28 ± 7 | 29 ± 9 | 0.3 |
| **Menopause** | 73 (89%) | 21 (84%) | 27 (87%) | 20 (95%) | 5 (100%) | 0.7 |
| **TSH, µIU/mL** | 1.61 ± 0.91 | 0.69 ± 0.19 | 1.44 ± 0.30 | 2.40 ± 0.24 | 3.94 ± 0.48 | <0.001 |
| **Levothyroxine dose, µg/day** | 93 ± 27 | 91 ± 30 | 97 ± 27 | 87 ± 21 | 95 ± 33 | 0.6 |
| **Vitamin B deficiency** | 7 (8.5%) | 2 (8.0%) | 2 (6.5%) | 3 (14%) | 0 (0%) | 0.8 |
| **Vitamin D deficiency** | 26 (32%) | 7 (28%) | 11 (35%) | 6 (29%) | 2 (40%) | 0.9 |
| **Major depressive disorder** | 18 (22%) | 4 (16%) | 7 (23%) | 6 (29%) | 1 (20%) | 0.8 |
| **Hypertension** | 60 (73%) | 20 (80%) | 22 (71%) | 13 (62%) | 5 (100%) | 0.3 |
| **Charlson Comorbidity Index** | 3.68 ± 2.19 | 3.36 ± 2.51 | 3.42 ± 2.05 | 4.14 ± 1.90 | 5.00 ± 2.35 | 0.2 |

<sup>1</sup> Mean ± SD; n (%)
<sup>2</sup> Kruskal-Wallis rank sum test; Fisher’s exact test

In the adjusted multivariable linear regression, higher TSH quartiles were associated with numerically greater ThyPRO symptom burden, although associations did not reach statistical significance (β=5.43, p=.216), Q3 (β=4.30, p=.389), and Q4 (β=3.76, p=.634). Major depressive disorder was independently associated with greater symptom burden (β=16.93, 95% CI 8.22–25.65; p<.001), as was hypertension (β=12.45, 95% CI 2.24–22.66; p=.018). No significant associations were observed for age, BMI, menopausal status, LT4 dosage, vitamin deficiencies, or CCI (Table 2). Ordered trend testing demonstrated no statistically significant monotonic increase in symptom burden across increasing TSH quartiles (Jonckheere–Terpstra JT=1219, p=.333).

**Table 2.** Associations between TSH quartiles and ThyPRO composite symptom burden using multivariable linear regression. Abbreviations: BMI - body mass index, CCI- Charlson Comorbidity Index, HTN- hypertension,
LT4 - levothyroxine, MDD- major depressive disorder.

| **Characteristic** | **Beta** | **95% CI** | **p-value** |
|---|---|---|---|
| TSH Quartile |  |  |  |
| Q1 | — | — |  |
| Q2 | 5.43 | -3.25, 14.11 | 0.216 |
| Q3 | 4.30 | -5.60, 14.20 | 0.389 |
| Q4 | 3.76 | -11.90, 19.42 | 0.634 |
| Age | -0.08 | -0.57, 0.41 | 0.748 |
| BMI | -0.47 | -1.06, 0.12 | 0.120 |
| Menopause | -3.01 | -17.23, 11.21 | 0.674 |
| LT4 dosage | 0.07 | -0.07, 0.21 | 0.312 |
| Vitamin B deficiency | 5.95 | -7.10, 19.00 | 0.366 |
| Vitamin D deficiency | -4.75 | -12.44, 2.95 | 0.223 |
| MDD | 16.93 | 8.22, 25.65 | <0.001 |
| HTN | 12.45 | 2.24, 22.66 | 0.018 |
| CCI | 0.54 | -1.89, 2.98 | 0.657 |

Abbreviation: CI = Confidence Interval

In exploratory domain-level analyses using continuous TSH, the strongest association was observed for the eye symptoms domain. Each 1 μIU/mL higher TSH was associated with a 9.57-point higher eye symptoms score (95% CI 3.12–16.03; raw p=.0049; Figure 2). This association did not remain statistically significant after Benjamini–Hochberg correction for multiple testing (false discovery rate-adjusted p=.058), and no individual ThyPRO domain remained statistically significant following false discovery rate correction. Continuous TSH was not significantly associated with prevalent hypertension (adjusted OR, 0.70; 95% CI, 0.34–1.45; p=.330), systolic blood pressure (β=1.42 mmHg per 1 μIU/mL higher TSH; 95% CI, −1.75 to 4.59; p=.375), or diastolic blood pressure (β=0.84 mmHg; 95% CI, −1.33 to 3.00; p=.445).

**Figure 2.**
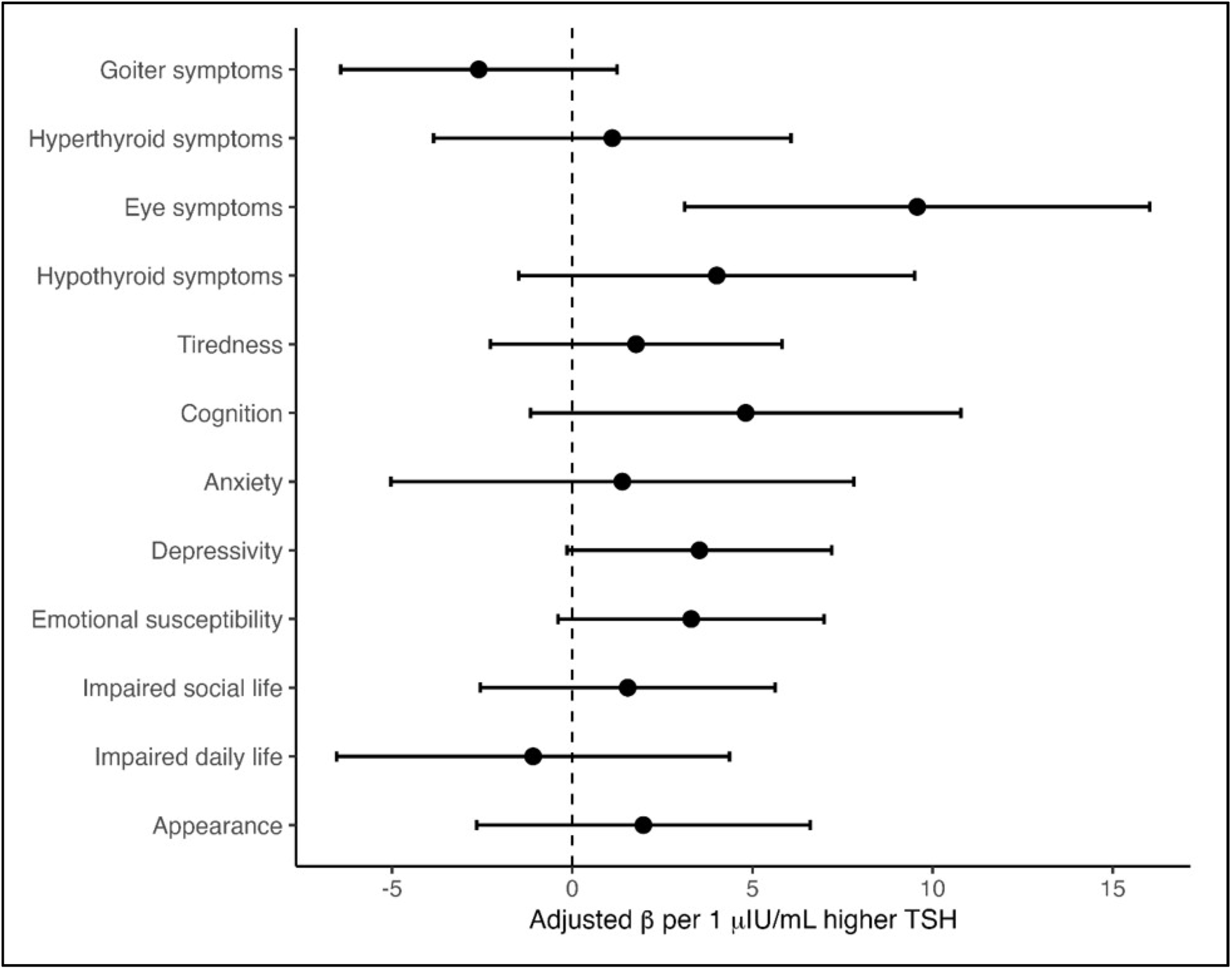
Exploratory domain-level analyses of specific symptom domains and serum thyroid-stimulating hormone levels in the absence of an association with overall symptom burden.

## Discussion and Conclusion

In this prospective cohort of women receiving stable LT4 therapy for overt hypothyroidism, serum TSH within the reference range was not independently associated with overall thyroid-related symptom burden as measured by the ThyPRO-39 instrument. Although mean composite scores were numerically higher in the upper TSH quartiles, neither multivariable regression nor ordered trend testing demonstrated a significant association between increasing TSH and overall symptom burden. Notably, MDD and HTN were independently associated with greater Thy-PRO-39 scores, suggesting that comorbidities may be more strongly related to persistent symptoms than modest variation in TSH within the euthyroid range.

This finding supports the clinical importance of comorbidity when interpreting persistent symptoms in treated hypothyroidism. Many complaints captured by this patient-reported outcome measure, including fatigue, impaired concentration, reduced activity tolerance, and mood symptoms, are not unique to thyroid dysfunction. The strong association with MDD highlights the potential contribution of overlapping symptom phenotypes. In contrast, the association with HTN may reflect the broader burden of cardiovascular disease and multimorbidity in this cohort. Recent data have associated suboptimal LT4 treatment and time spent outside the therapeutic TSH range with modest increases in mean blood pressure and blood pressure variability.^15^ However, in our biochemically euthyroid cohort, exploratory analyses did not demonstrate significant associations between continuous TSH and prevalent hypertension, systolic blood pressure, or diastolic blood pressure. Persistent symptom burden may therefore reflect overlapping disease processes rather than incomplete biochemical thyroid control alone.^16,17^

The absence of statistically significant findings should be interpreted within the context of several important considerations. First, symptom burden demonstrated substantial heterogeneity across participants, particularly within higher TSH quartiles. Such variability is common in patient-reported outcome research and likely reflects the multifactorial nature of symptom experience among these individuals. Additionally, the highest TSH quartile contained only five participants, limiting statistical precision and contributing to wider confidence intervals. As a result, the modest cohort size may therefore have limited the ability to detect a clinically relevant pattern.

Exploratory domain-level analyses were performed to determine whether TSH was associated with specific symptom domains despite the absence of an association with overall symptom burden. When TSH was analyzed as a continuous measure, the strongest association was observed for the eye symptoms domain, with higher TSH associated with greater burden of eye symptoms such as grittiness, impaired vision, and light sensitivity (Figure 2). Although no individual domain remained statistically significant after correction, the observed heterogeneity highlights the complexity of symptom experience and supports continued investigation of symptom-specific phenotypes. Such approaches may become increasingly important as thyroid disease management evolves toward more personalized and precision health-oriented models of care.

This study has several strengths, including contemporaneous TSH measurement at survey administration, use of a validated thyroid-specific QoL instrument, and multivariable analyses accounting for clinically relevant covariates. Several limitations should also be considered. The study cohort was modest in size and had unequal quartile distributions, particularly in Q4, limiting precision and power to detect smaller associations. Additionally, participants were recruited from a Women’s Heart Center, providing a clinically relevant perspective on symptom burden among women with substantial cardiovascular comorbidity but potentially limiting generalizability to men, patients managed exclusively in endocrinology settings, or broader primary care populations. As an observational study, residual confounding and causal inference also remain important limitations.

These findings contribute to an evolving understanding of symptom burden in treated hypothyroidism. Evaluating patient-reported outcomes alongside biochemical markers may support a more patient-centered approach by recognizing that persistent symptoms can reflect factors beyond serum TSH alone. Our results suggest that comorbidities may be particularly important when interpreting residual symptoms in biochemically euthyroid patients. Future studies incorporating larger, more diverse cohorts, longitudinal thyroid measurements, and broader clinical characterization will help refine these approaches and optimize treatment strategies within a precision health framework.

## Data Availability

Data is available to be viewed if requested.

## Acknowledgments

None

## Abbreviations

BMI: body mass index
CCI: Charlson Comorbidity Index
HTN: hypertension
LT4: levothyroxine
MDD: major depressive disorder
QoL: quality of life
ThyPRO: Thyroid-Related Patient-Reported Outcome
TSH: serum thyroid-stimulating hormone

## Supplemental Figure

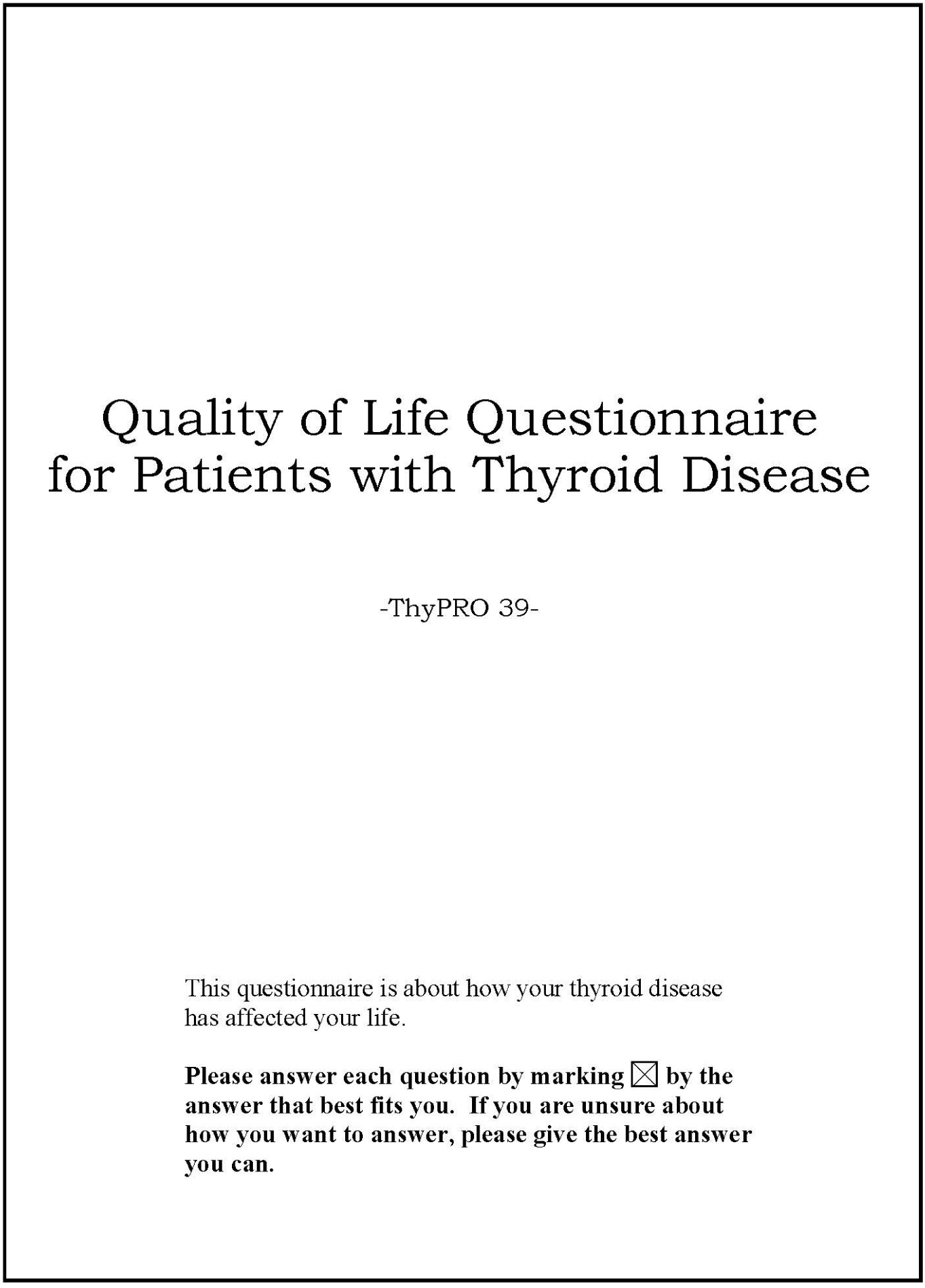

The first section of the questionnaire is about symptoms, tiredness, memory, mood, and health.

Please base your **answers on how you have been feeling <u>in general</u>** during the past 4 weeks.

1. ***The first questions are about symptoms***

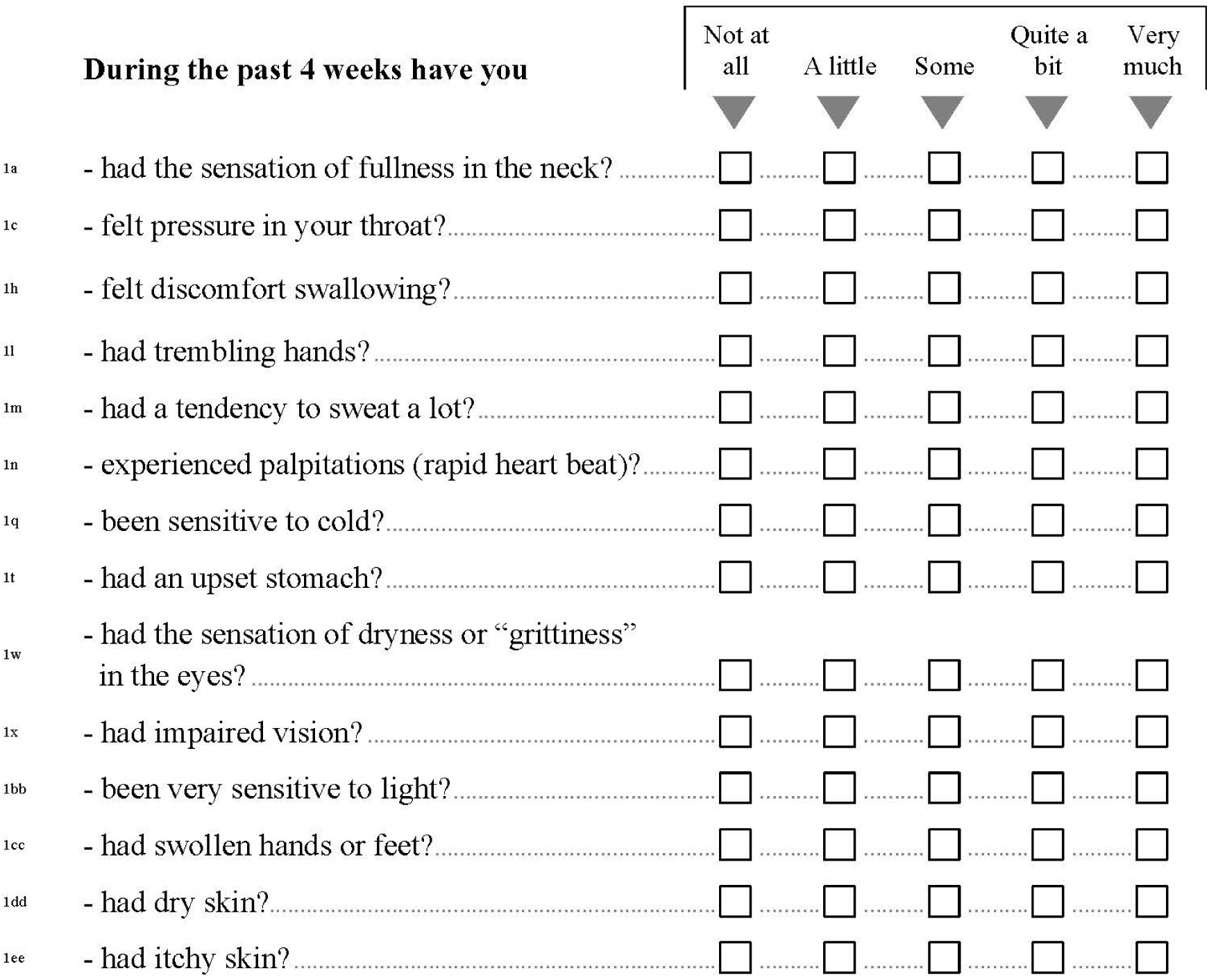
2. ***The following question are about triedness***

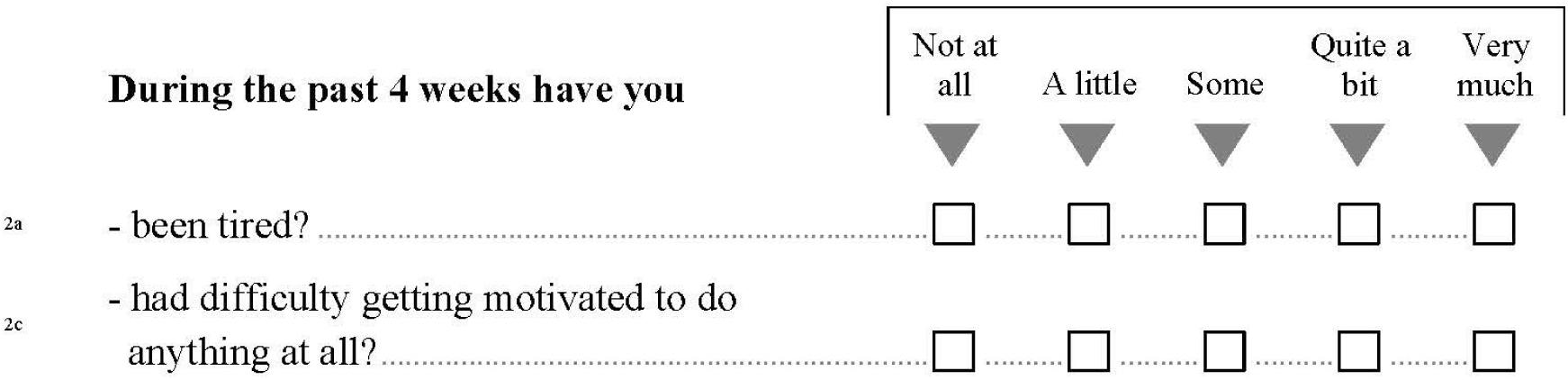
3. ***The following question is about your vitality***

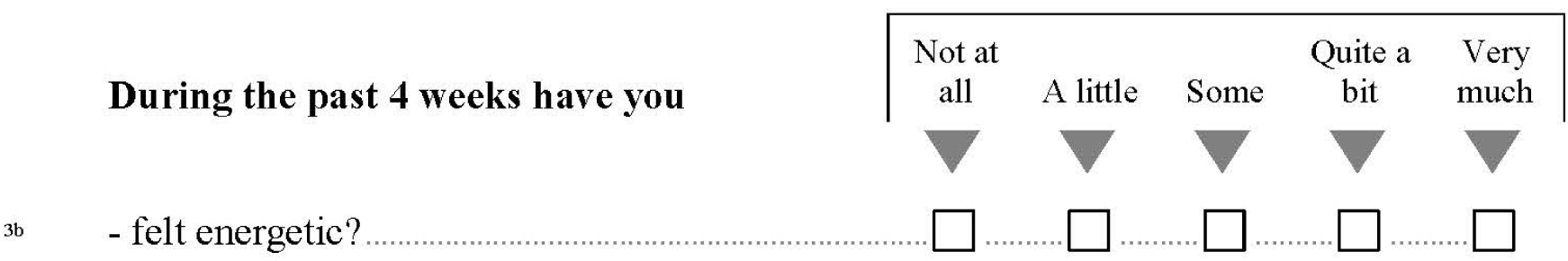
4. ***The following question are about memory and concentration***

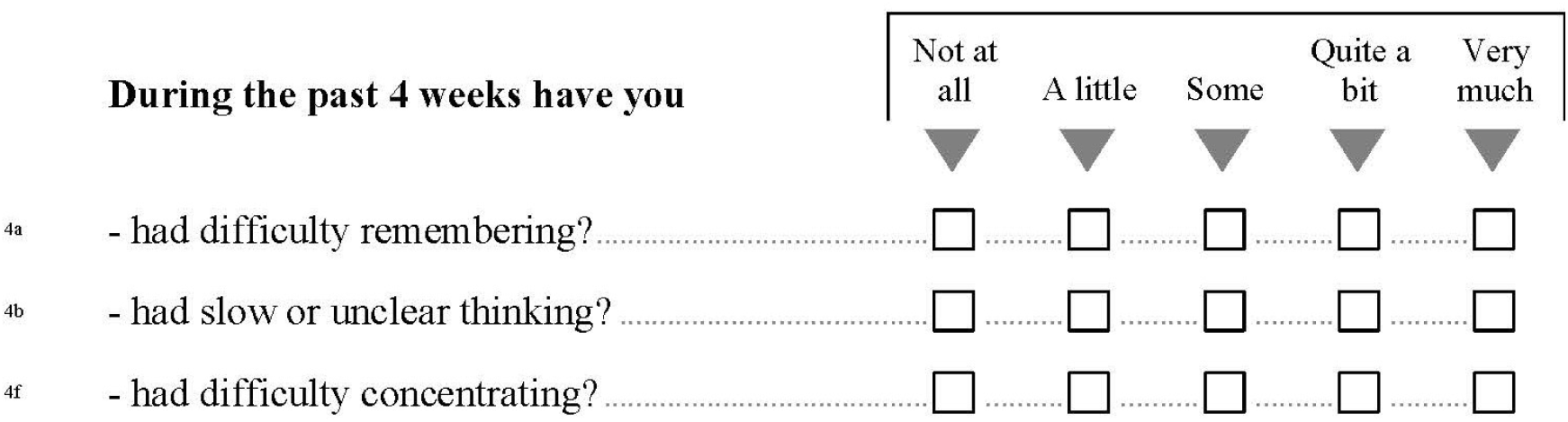
5. ***The following questions are about nervousness and tension***

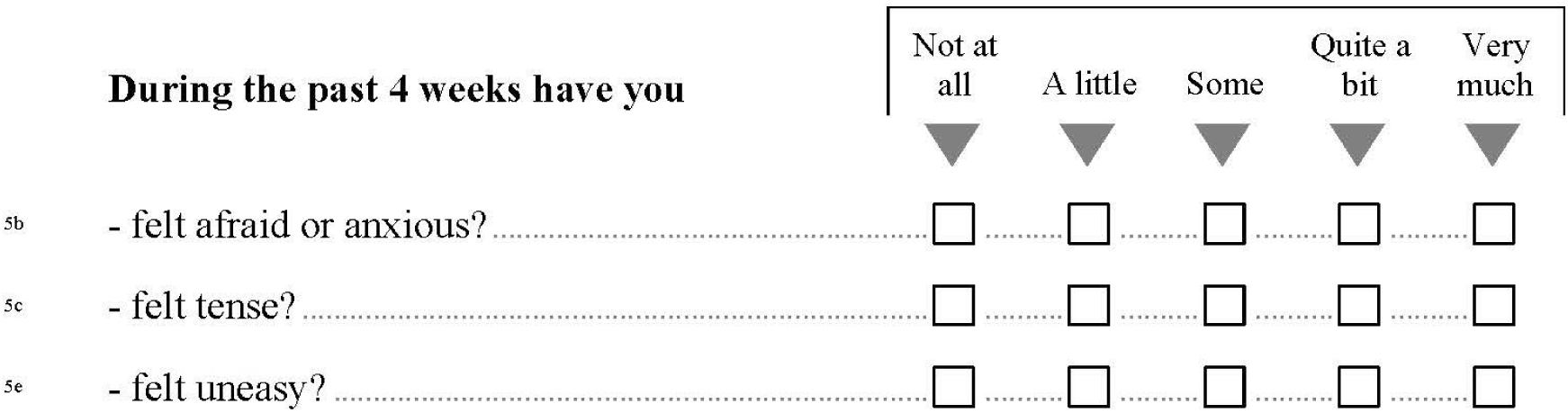
6. ***The following questions are about psychological well-being***

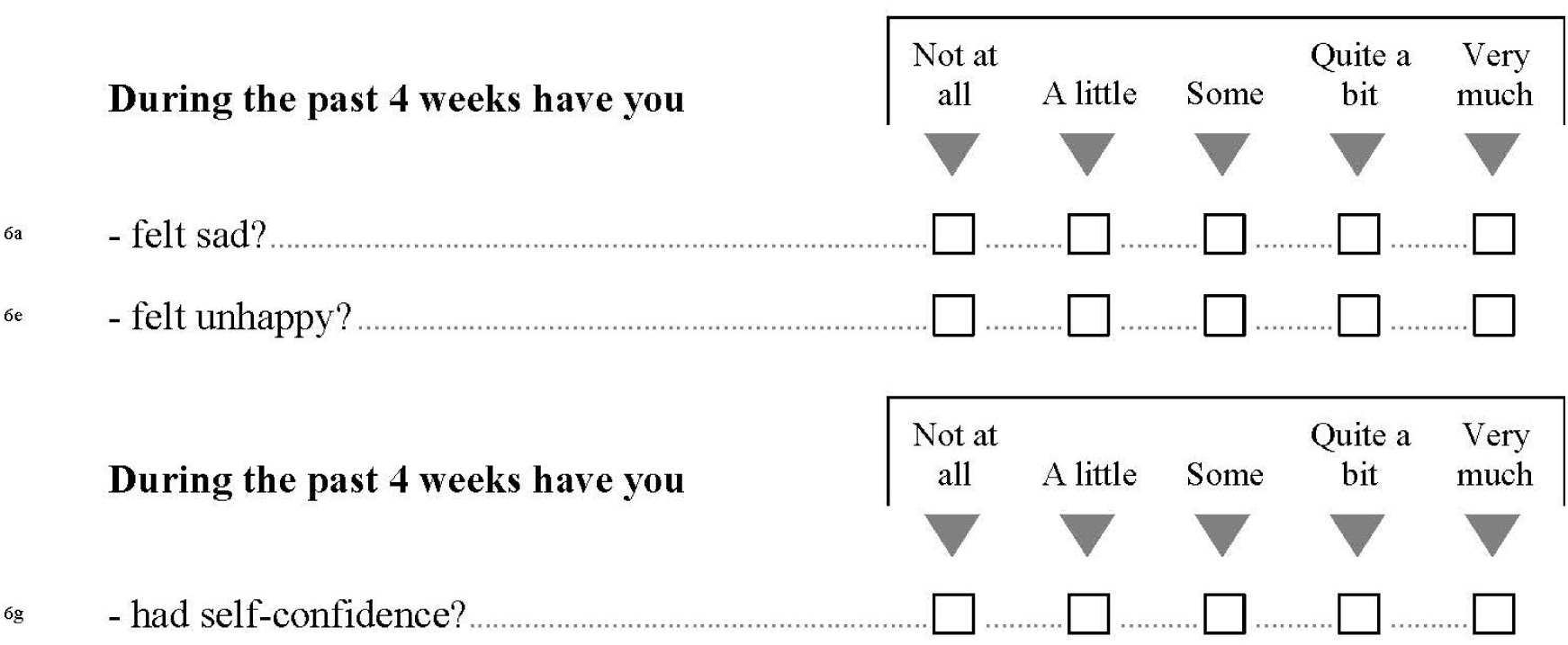
7. ***The following questions are about having difficulty coping or having mood swings***

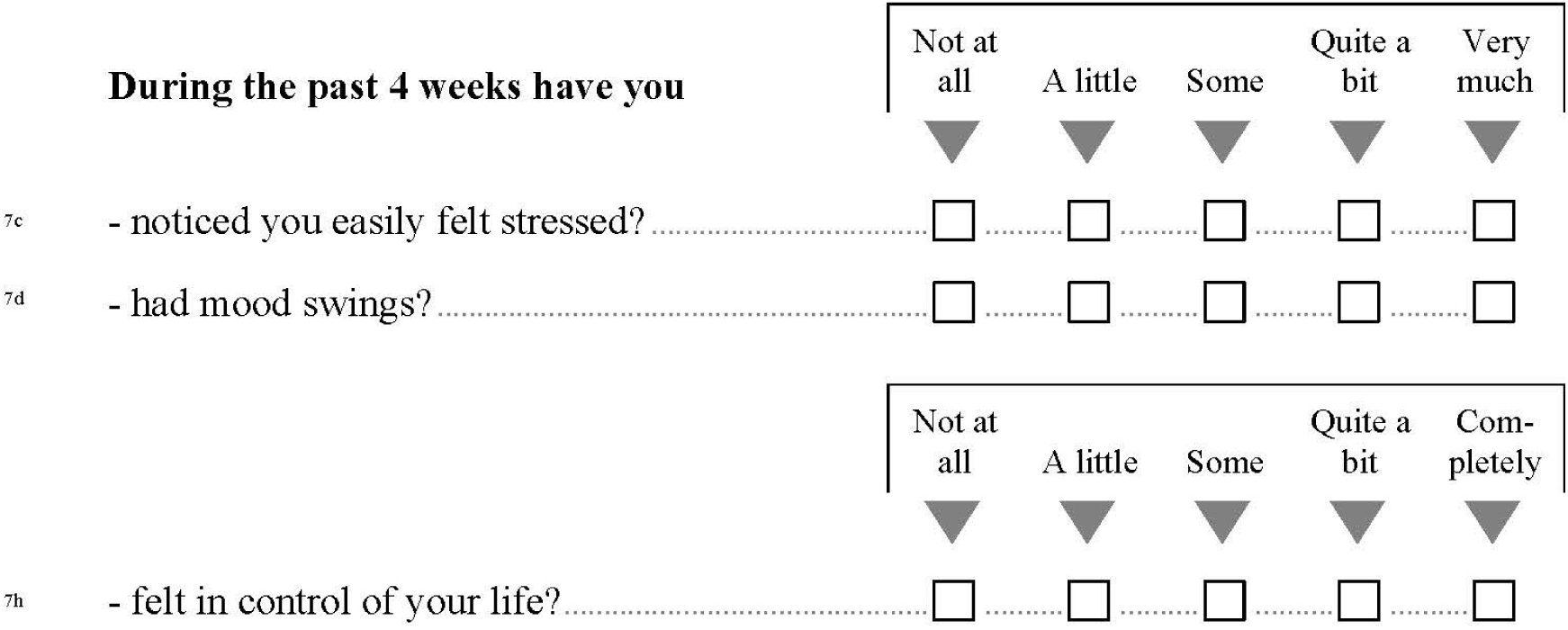

The remainder of the questionnaire is about **how your thyroid disease may have affected various aspects of your life**
8. ***The following questions are about your relationships with other people***

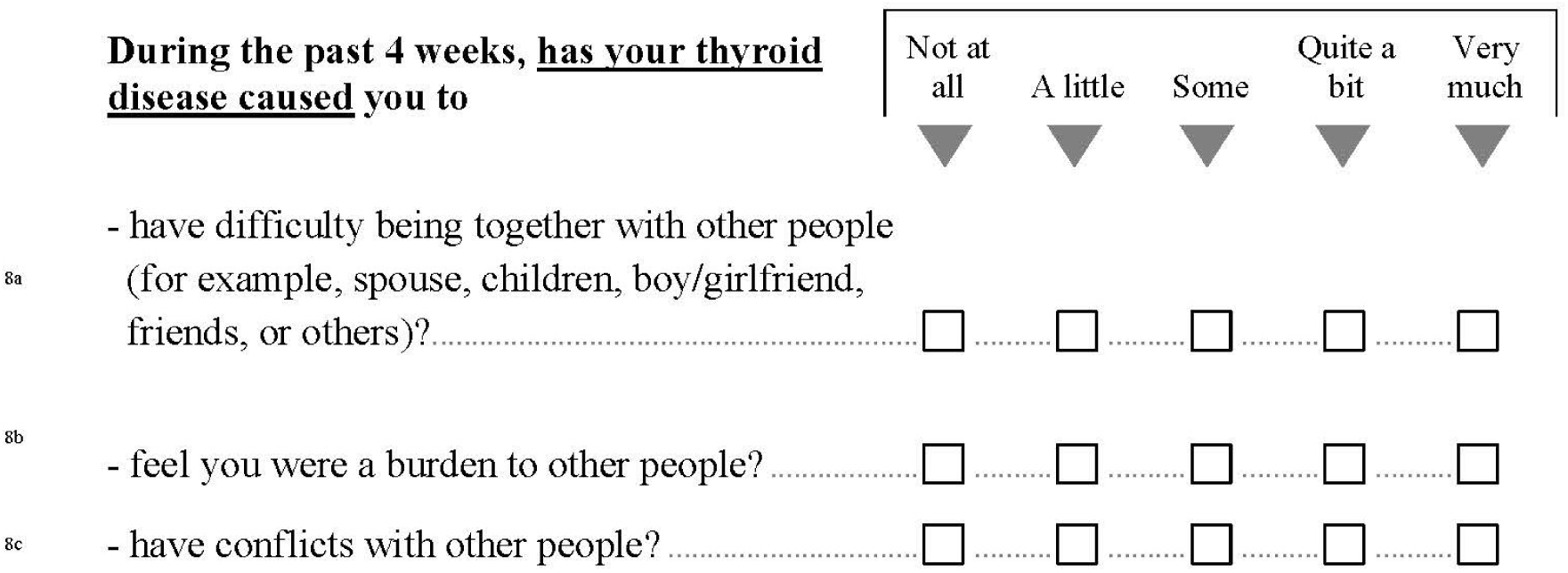
9. ***The following questions are about your daily activities***

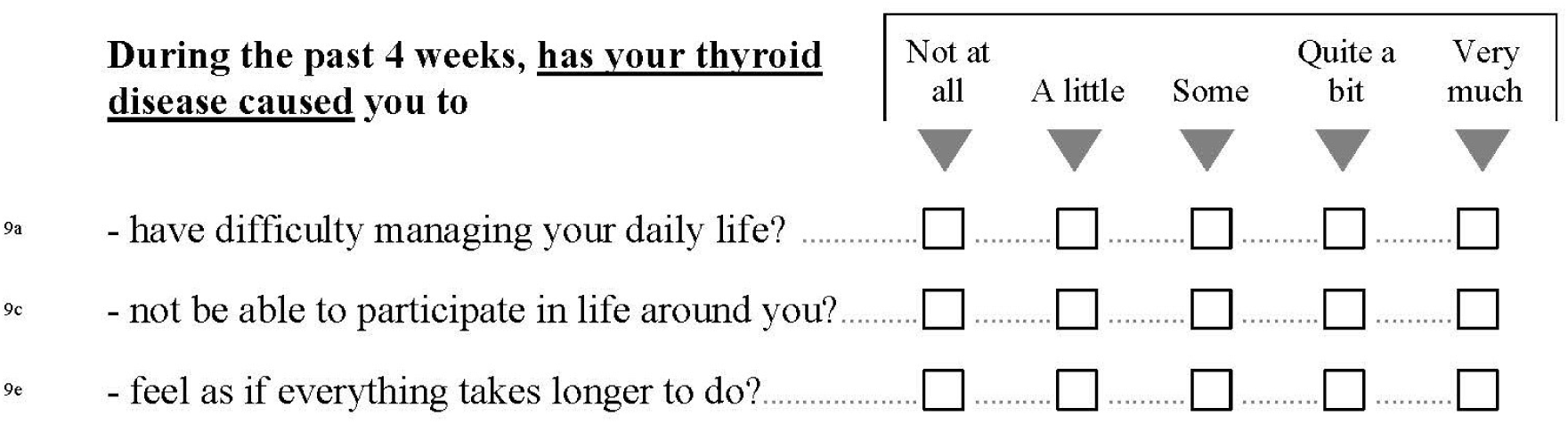
10. ***Thyroid diseases (or their treatment) may affect your <u>appearance.</u> (For example, by causing swelling of the neck, swollen face, hands, or feet, or changes in weight or to the eyes.)***

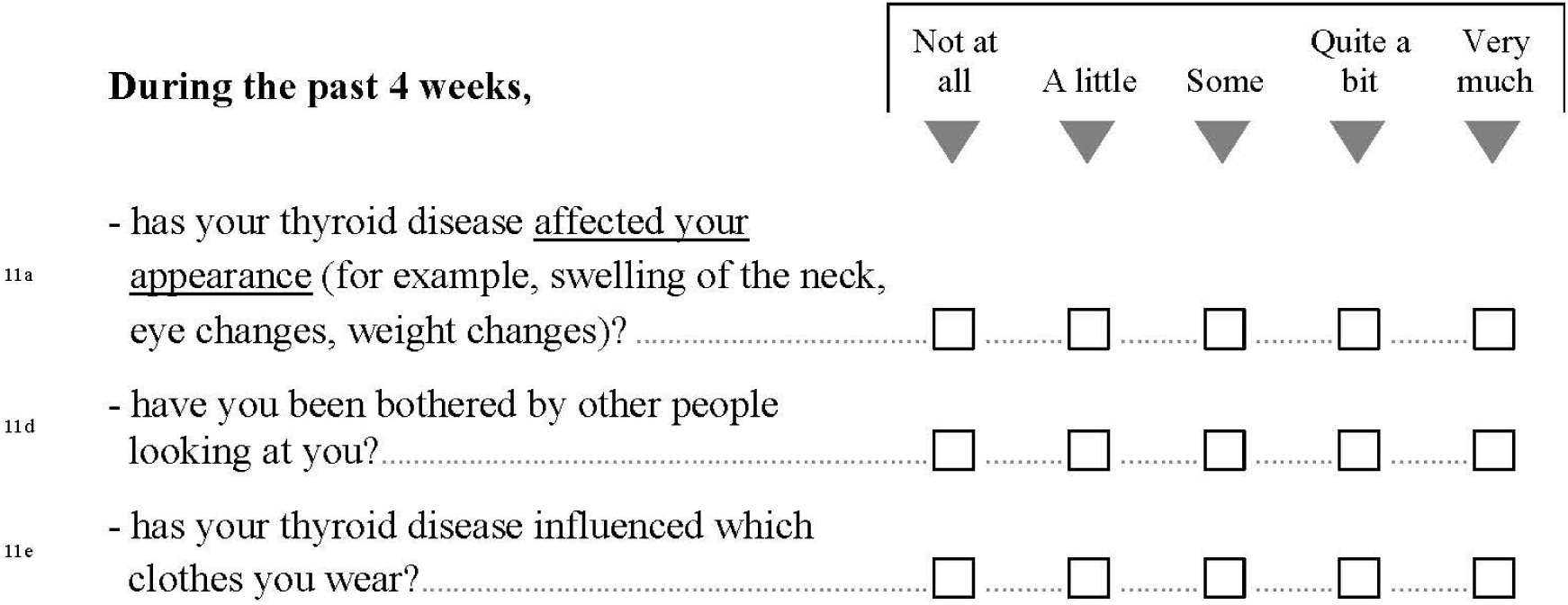

**12. *The final question is about to what extent your thyroid disease has affected you overall during the past 4 weeks***

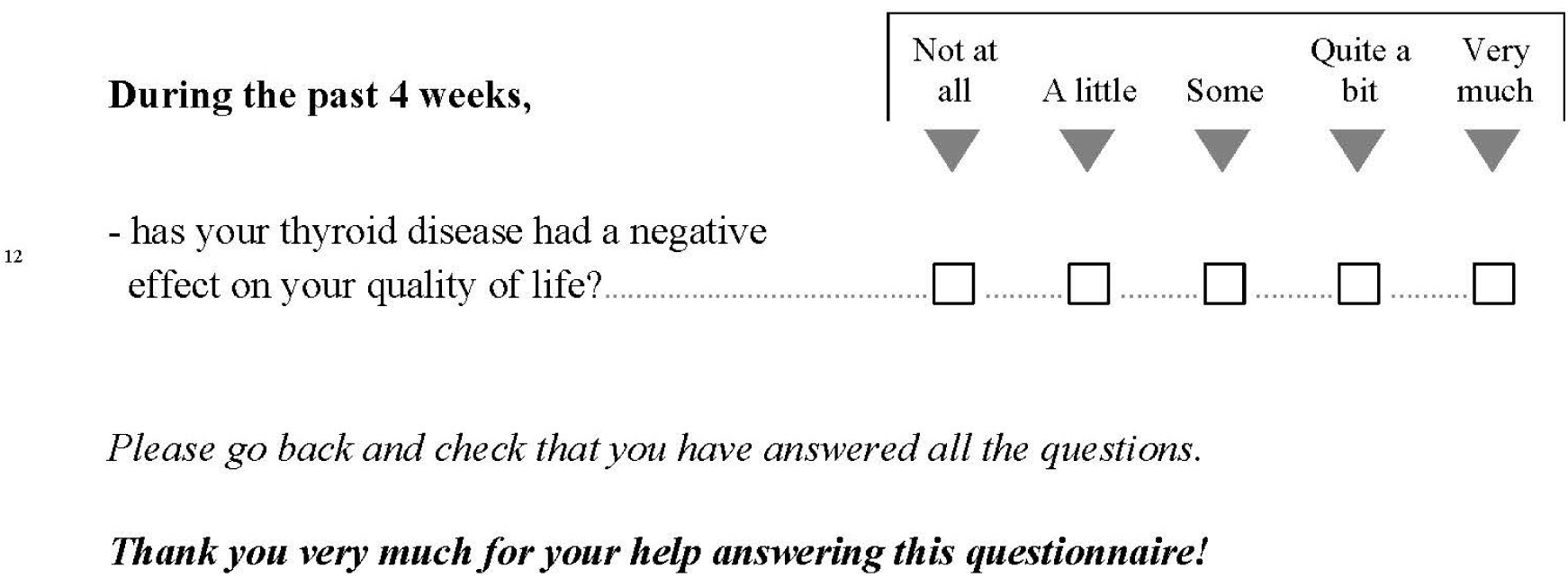

